# Loss of filtration function in diabetic glomeruli is associated with ultrastructural changes in glomerular endothelial cell fenestrations

**DOI:** 10.1101/2020.12.15.20248257

**Authors:** Natalie C Finch, Sarah S Fawaz, Chris R Neal, Mathew Butler, Vivian Lee, Abigail Lay, Megan Stevens, Lusyan Dayalan, Hamid Band, Harry Mellor, Gavin Welsh, Rebecca R Foster, Simon C Satchell

## Abstract

**Background:** The role of glomerular endothelial cell fenestrations in renal filtration function and development fenestral ultrastructural changes in disease contributing to loss of function is poorly described.

**Methods:** We determined glomerular ultrastructural changes with comprehensive characterisation of fenestral findings and also functional changes including measurement of glomerular ultrafiltration coefficient in diabetic mice and humans. We further evaluated Eps homology domain protein 3 (Ehd3) as a potential regulator of glomerular endothelial cell fenestrations.

**Results:** This study identified loss of GEnC fenestration density which was associated with decreased glomerular ultrafiltration coefficient and GFR in diabetic nephropathy. We also identified increased GEnC fenestration width, an ultrastructural change that may develop to maintain filtration surface area. GEnC fenestration width was negatively associated with glomerular ultrafiltration coefficient and GFR considered to be a result of the development of diaphragms in widening fenestrations providing resistance to filtration. The increased presence of diaphragmed fenestrations in diabetes was supported by increased PV1 expression. We further identified decreased glomerular Ehd3 expression in diabetes and demonstrated its association with GEnC fenestration measurements suggesting its role in regulating fenestrations. We also demonstrated its positive associations with filtration function suggesting loss of glomerular Ehd3 expression in disease may contribute to declining glomerular filtration function through aberrant GEnC fenestration regulation.

**Conclusions:** This is the first study to demonstrate the critical role of glomerular endothelial cell fenestrations in renal filtration function and identify a potential key regulator that may serve as a therapeutic target to retore filtration function in disease.

## Introduction

Glomerular endothelial cells (GEnC) are highly differentiated endothelial cells that line the glomerular capillaries in the kidney. They are perforated with transcytoplasmic pores, reportedly 60-80nm in diameter,^1^ known as fenestrations. GEnC fenestrations allow the passage of fluid and small solutes from the glomerular capillary lumen through the endothelial cells, without the need for endocytosis or receptor mediated mechanisms. The GEnC fenestrations are considered empty pores apart from the presence of the endothelial glycocalyx at the periphery. Quiescent GEnC have predominantly non-diaphragmed fenestrations^1^ and the lack of a structural barrier enables high fluid flux driven by hydrostatic pressure. Alterations in GEnC fenestration density or diameter pose an important potential mechanism for regulating glomerular filtration and may play a critical role in pathogenesis of diseases characterised by a decline in glomerular filtration rate (GFR).

The glomerular ultrafiltration coefficient (L_P_A) can be determined in isolated glomeruli using an oncometric assay.^2-5^ This established method allows the evaluation of hydraulic permeability of the glomerular filtration barrier in isolation from circulating and haemodynamic factors. It is a measurement of the filtration in an individual glomerulus in contrast to GFR which determines whole kidney filtration function. Thus, measurement of glomerular ultrafiltration coefficient allows its relationship with ultrastructural components of the glomerular filtration barrier to be evaluated. The contribution of GEnC fenestrations to the glomerular ultrafiltration coefficient is relatively unstudied. This is potentially related to early mathematical models suggesting that the glomerular endothelium provides little resistance to hydraulic permeability.^6^ However, subsequent studies have demonstrated that GEnC fenestration density contributes to decreased glomerular hydraulic permeability in women with pre-eclampsia.^7^ Further, decreased glomerular ultrafiltration coefficient has been demonstrated in VEGF_165_b overexpressing mice with decreased GEnC fenestration density.^5^ Decreased glomerular ultrafiltration coefficient may, in combination with haemodynamic changes, result in decreased glomerular perfusion which in combination with loss of nephron number contribute to declining GFR. To the author’s knowledge, relationships between GEnC fenestration measurements and *ex vivo* measurements of glomerular ultrafiltration coefficient, as opposed to mathematical modelling, and GFR have not previously been described.

Diabetic nephropathy is the leading cause of end stage renal disease in humans. It is characterised by glomerular hypertrophy and hyperfiltration in early stage disease and progressive albuminuria, decline in GFR and glomerular and tubulointerstitial structural changes in later stage disease.^8^ Ultrastructural changes in the glomerulus associated with the later stages of diabetic nephropathy, in people in which filtration function has declined, include loss of GEnC fenestrations.^9^ Decreased percentage of fenestrated endothelium is correlated with GFR in people with type 2 diabetes^9, 10^ and also reported in people with type 1 diabetes.^11^ BTBR *ob*^-^*/ob*^-^ mice develop type 2 diabetes and progressive diabetic nephropathy.^12^ Their unique susceptibility to diabetic nephropathy has been suggested to be related to endothelial dysfunction similar to that seen in eNOS deficient mice.^12^ However, the GEnC fenestral ultrastructural changes have not been described.

Knowledge of factors that regulate GEnC fenestrations will allow the development of targeted therapies to manipulate them and potentially improve glomerular filtration function in disease. Angiogenic factors such as VEGF-A are known regulators of GEnC fenestrations. Eps15 homology domain-containing protein 3 and 4 (Ehd3 and 4) are endosomal transport proteins.^13^ Within the kidney, Ehd3 is specifically expressed in GEnC^14, 15^ and has been localised to the fenestrae.^14^ Ehd4 is highly expressed in peritubular endothelial cells with very low expression in GEnC under normal conditions.^16^ Dual knockout of Ehd3 and 4 in mice resulted in development of lesions consistent with thrombotic microangiopathy with absence of GEnC fenestrations.^16^ This suggests a critical role for Ehd3 in maintaining a healthy GEnC phenotype and as candidate regulator of GEnC fenestrations.

In this study, we aim to examine the role of GEnC fenestrations in filtration function at the glomerular filtration barrier in diabetes analysing samples from both the BTBR *ob/ob* mouse model and human patients. We further aim to study Ehd3 glomerular expression in diabetes and its association with GEnC fenestration measurements and glomerular filtration function and demonstrate its role as a key regulator of GEnC fenestrations. We hypothesise that GEnC fenestral ultrastructural changes develop in diabetes and contribute to loss of glomerular ultrafiltration coefficient and GFR. In addition, that glomerular Ehd3 expression decreases in diabetes and correlates with GEnC fenestration measurements and glomerular filtration function and *in vitro* loss of Ehd3 results in decreased fenestration formation.

## Methods

### Mice

Male wild type (+/+) and homozygous BTBR *ob*^-^*/ob*^-^ mice were obtained from the Jackson laboratory (BTBR.Cg-*Lep*^*ob*^/WiscJ; Bar Harbor, ME, USA). The mice were provided with food and water *ad libitum* and maintained in a clean temperature-controlled environment under a 12-hour light/dark cycle. Animal experiments were conducted with approval by the UK Home Office and in accordance with the Animals (Scientific Procedures) Act 1986. Weekly measurement of body weight and bi-weekly blood glucose measurement via tail vein blood collection was performed. At 20 weeks of age, mice underwent terminal anaesthesia via intraperitoneal injection of pentobarbitone (Euthatal 200mg/ml) and both kidneys were harvested and immediately processed as described in the following methods.

### Renal functional measurements

#### Urinary albumin to creatinine ratio

Spot urine samples were collected bi-weekly. Mice were placed in a metabolic cage for up to 2 hours. Terminal urinary collection was performed via cystocentesis. Urinary albumin concentration was quantified using a mouse albumin ELISA (Bethyl Laboratories Inc; Montgomery, TX) and urinary creatinine concentration determined at a commercial reference laboratory (Langford Vets Diagnostic Laboratories, Bristol, UK) using an enzymatic spectrophotometric assay (Konelab T-Series 9812845; Thermo Fisher Scientific, Vantaa, Finland). Urinary albumin to creatinine ratio (uACR) was calculated as urinary albumin concentration/ urinary creatinine concentration.

#### Glomerular ultrafiltration coefficient (L_P_A/V_i_)

Glomeruli were isolated from freshly harvested kidney tissue using a standard sieving technique. Glomerular ultrafiltration coefficient was determined in individual glomeruli *ex vivo* within 4 hours of isolation, using a previously described oncometric method, and normalised to glomerular volume (L_P_A/V_i_).^2-5^ Between 3 and 12 glomeruli per mouse were analysed using image analysis software (FIJI) by a blinded investigator.

#### Glomerular filtration rate

Glomerular filtration rate (GFR) was determined by measuring endogenous creatinine clearance. Within 24 hours of terminal anaesthesia mice were placed in a metabolic cage and timed urine collection performed over approximately six hours with exact time recorded in minutes. Plasma creatinine concentration was measured in a terminal blood collection sample collected into a heparinised plasma tube. Plasma and urinary creatinine concentration were determined as described above. Endogenous creatinine clearance was calculated using the standard equation GFR (mL/min) = (U_Cr_ x V)/ P_Cr_ x T) where U_Cr_ is urinary creatinine concentration (µmol/l), V is volume (ml), P_Cr_ is plasma creatinine concentration (µmol/l) and T is time (mins).

Transmission electron microscopy to determined glomerular ultrastructural measurements Immediately following terminal anaesthesia, kidney tissue was harvested and 1mm^3^ diced kidney cortex obtained and transferred to 2.5% glutaraldehyde in 0.1 M cacodylate buffer. Samples were post-fixed in 1% osmium tetroxide followed by ethanol dehydration and embedding in TAAB 812 resin (Agar Scientific). Sections were cut at 50-100nm using an ultramicrotome and stained with 3% aqueous uranyl acetate followed by Reynolds’ lead citrate. Electron micrographs were acquired using a Technai 12 electron microscope (FEI, Hillsboro, Oregan). Image acquisition was performed in a standardised manner by acquiring images of 3 glomeruli /mouse with six random images per glomerulus, 3 at 12 o’clock and 3 at 6 o’clock position from the low power whole glomerulus image. Additional kidney tissue from BTBR *ob/ob* mice aged 6, 10, 15 and 20 weeks was contributed by University College London (David Shima) in 2.5% glutaraldehyde in 0.1 M cacodylate buffer and stored and shipped at 4°C. Samples were processed as described above. Image analysis was performed by a blinded investigator using image analysis software (FIJI). GEnC fenestration density was determined by counting the number of fenestrations per unit length of the GEnC peripheral cytoplasm. Glomerular endothelial cell fenestration width was determined by measuring the diameter of the fenestration at the narrowest distance between the opposing cell membranes. Diaphragms were identified as a single clear line of electron dense material spanning the fenestration and determined as the percentage of the total number of fenestrations. GEnC filtration surface area was calculated as a percentage of measured GEnC surface area covered by total fenestration widths. In addition, podocyte slit density, slit width, foot process width and glomerular basement membrane thickness were determined. Mean values per individual were used for statistical analysis.

### PLVAP immunofluorescence to determine glomerular expression

Freshly harvested renal cortical tissue was immediately transferred to liquid nitrogen and stored at -80°C. Frozen sections (4µm) of cortical tissue were cut at the University of Bristol Histology Service. Sections were briefly fixed in 4% (wt/vol) PFA followed by a blocking step (1% BSA in PBS). Primary antibody (Table 1) was applied to sections overnight. The following day sections were incubated with secondary antibody (1:200 anti-rat 488 Alexa Fluor, Life Technologies, Thermo Fisher Scientific) and the nuclei were counterstained with 4’,6-diamidino-2-phenylindole (Invitrogen, Thermo Fisher Scientific). Sections were mounted in Vectashield mounting medium (Vector Laboratories). Image analysis was performed using an image software program (FIJI) by a blinded investigator. Corrected total glomerular fluorescence intensity was determined in three glomeruli per mouse and the mean for each mouse used in statistical analyses. Images were obtained at x40 magnification using an AF600 LX wide-field fluorescence microscope (Leica Microsystems, Milton Keynes, UK).

**Table 1:** List of antibodies

| Antibody | Host species | Dilution | Manufacturer and product number | Samples applied to |
| --- | --- | --- | --- | --- |
| Ehd3 | Rabbit | 1:100 | Kindly gifted from Dr Hamid Band, University of Nebraska | Mouse, b.End5 |
| Ehd4 | Rabbit | 1:100 | Kindly gifted from Dr Hamid Band, University of Nebraska | Mouse |
| Ehd 3 | Rabbit | 1:50 | Novus NBP2-31894 | Human |
| Ehd 4 | Rabbit | 1:50 | Proteintech 11382-2-AP | Human |
| PV-1 | Rat | 1:50 | Santa Cruz sc-19603 | Mouse |

### Ehd 3 and 4 immunohistochemistry to determine glomerular expression profiles

Freshly harvested cortical renal tissue was transferred to 4% (wt/vol) paraformaldehyde (PFA) for 48hrs hours. Dehydration and paraffin embedding were performed at the University of Bristol Histology Service. Sections (4µm) were cut using a microtome and transferred to glass slides. Deparaffinisation and hydration steps involved incubation in Histoclear II and decreasing concentrations of ethanol. Antigen retrieval was performed by heating sections in 10mM sodium citrate buffer (pH 6). Non-specific IgG binding was blocked with 1% BSA and 10% normal goat serum in TBS-Triton-X (0.1%). Primary antibodies (Table 1) or an IgG control were applied to sections overnight. The following day endogenous peroxidase activity was blocked with 3% (wt/vol) hydrogen peroxide and sections incubated with an HRP conjugated secondary antibody specific for the antibody (SignalStain Boost IHC Detection Reagent, Cell Signaling, Danvers, MA, USA). Sections were subsequently incubated with DAB substrate (SignalStain DAB substrate, Cell Signaling, Danvers, MA, USA) until sections changed colour from purple to brown. Sections were counter-stained with haematoxylin. Image analysis was performed using an image software program (FIJI) by a blinded investigator. Corrected total staining intensity was determined in three capillaries per glomeruli and three glomeruli per mouse and the mean for each mouse used in statistical analyses. Images were obtained at x40 magnification using light microscopy.

RNA extraction and qPCR to determine glomerular PLVAP, Ehd3 and Ehd4 mRNA expression. Freshly sieved glomeruli were obtained from renal cortical sections by passing tissue through sequential sieves to extract the glomeruli and were immediately transferred to storage at -80°C prior to RNA extraction. Glomerular RNA extraction was performed using the Qiagen RNEasy kit (cat. no. 74104). Glomeruli were drawn repeatedly into a 0.5ml syringe to aid in cellular lysis. Following lysis, measurements of RNA concentrations were obtained using a nanophotometer (Pearl Implen, München, Germany) prior to cDNA conversion. RNA was converted to cDNA using a high capacity RNA-to-cDNA kit (ref 4387406; Applied Biosystems, Foster City, California, USA). The primers were designed using Eurofins; PLVAP Forward 5’ CTATCATCCTGAGCGAGAAGC 3’, Reverse 5’ GCAGCAGGGTTGACTACAGG 3’; Ehd3 Forward 5’ CGCCGTGCTTGAAAGTATCAG 3’, Reverse 5’ ATAATTCGGTCCACCCGCTC 3’; Ehd4 Forward 5’ ACCAAGTTCCACTCACTGAA 3’, Reverse 5’ GTTCATCTCCTCCTGGCTGA 3’. Optimum primer concentrations were established and a standard protocol used for quantitative polymerase chain reaction (qPCR) analysis using the Fast Sybr Green master mix (ref 438612; Applied Biosystems). Statistical analyses were performed on delta cycle threshold values and data are shown as fold-change + SD.

### Human samples

All studies on human kidney tissue were approved by national and local research ethics committees (REC) and conducted in accordance with the tenets of the Declaration of Helsinki. Transmission electron microscopy images and renal biopsy samples were obtained from Bristol, UK (Histopathology Department, Southmead Hospital) and were archived anonymously (REC H0102/45). Tissue was immersion fixed in 10% neutral buffered formalin and embedded in paraffin. Samples had been clinicopathologically diagnosed and included patients with diabetic nephropathy and patients with thin basement membrane nephropathy which served as a control population. Patient eGFR, where available, was also provided. Transmission electron micrograph image analysis was performed as described above. In addition, rejected kidney transplant tissue was also obtained and included in the control population for immunohistochemistry studies. Immunohistochemistry was performed as described above. Gene expression data were extracted from the Nephroseq database (www.nephroseq.org).

### *In Vitro* fenestration formation

A mouse brain endothelioma cell line (b.End5) was obtained from Culture Collections, Public Health England, Porton Down, UK) and maintained in high glucose DMEM (Sigma-Aldrich, Gillingham, UK) containing 10% FBS. An Ehd3 knockdown and control scrambled b.End5 cell line were generated. The lentiviral vectors containing mouse Ehd3 shRNA or scrambled sequences were purchased from Dharmacon Horizon Inspired cell solutions (VGH5526-EG57440). b.End5 at 40-60% confluency was incubated with the lentiviral particle in the presence of polybrene at 1:100 ratio for 4h in serum free media. The infected cells were cultured in complete media for 48h, followed by a puromycin selection at 0.8µg/ml for 3 consecutive days to obtain stable knockdown cell lines. The knockdown efficiency was confirmed by qPCR and Western blotting. Briefly, RNA was extracted from b.End5 cells (scrambled and knockdown) and PCR performed using the protocol described above. For Western blot analysis, b.End5 cells (scrambled and knockdown) were washed with PBS prior to protein extraction. The cells were lysed with ice-cold RIPA buffer (ThermoFisher Scientific # 89900) followed by centrifugation of lysates at 13000 rpm for 15 min at 4°C and collection of supernatant. Supernatant was added to Laemmli sample buffer at a 1:4 sample volume ratio, denatured in a heat block at 90°C for 10 minutes, separated on a 10% SDS-polyacrylamide gel and transferred to a polyvinylidene difluoride membrane. Following a blocking step (3% BSA prepared in Tris-buffered saline-tween (TBST; 20 nM Tris (pH 7.2), 150 mM NaCl, 0.1% Tween 20) for 1h, immunoblots were incubated overnight at 4 °C with primary antibodies to detect Ehd3 (Table 1). Housekeeping gene β-actin (1:5000; Millipore, Billerica, MA, USA) was used for normalization. Blots were incubated with secondary antibody for 1h at room temperature. Luminal and Femto peroxidase (Western ECL Substrate, Biorad Clarity) were added in equal volumes (500µl each) to the membrane and the signal analysed using an Amersham imager 600 system. Densitometry was performed using ImageJ 1.43m software.

b.End5 cells were seeded at a density equivalent to 1.5 x 10^6^ cells per 100mm dish onto gold grids on glass coverslips coated with pioloform. After 24hrs, fenestrations were induced with 1.25µM Latrunculin A (Sigma-Aldrich, Gillingham, UK) for 3hrs or 100ng/ml mouse VEGF_164_ (R&D Systems, Minneapolis, USA) for 24hrs. Cells were fixed in 2.5% glutaraldehyde in 0.1 M cacodylate buffer, post-fixed in 1% osmium tetroxide followed by ethanol dehydration and dried using critical point drying. Wholemount cell samples were imaged using a Technai 12 electron microscope (FEI, Hillsboro, Oregan). Between 8 and 9 cells per treatment group were analysed. Fenestration density was determined in a 720nm^2^ area in sieve plates in two separate areas per cell. Fenestration width was measured for each individual fenestration in a 720nm^2^ area. Mean fenestration density and width per cell was used for statistical analyses.

### Statistical analyses

Statistical analysis was performed using GraphPad Prism (version 8, La Jolla, CA, USA). Gaussian distribution was demonstrated, and parametric statistical testing performed. Data are expressed as mean ± standard deviation (SD). Mean values per individual were used for statistical analysis where multiple measurements per individual were obtained. Across group comparisons were performed using Student’s *t* test and one-way ANOVA. Post-hoc analysis was performed using Tukey’s multiple comparisons test. Relationships between variables were evaluated by performing linear regression and determining the coefficient of determination (R squared, r^2^) and correlations were evaluated by determining Pearson’s correlation coefficient (r). Significance was set at P <0.05.

## Results

### Diabetic nephropathy in BTBR *ob*^*-*^*/ob*^*-*^ mice is associated with decreased fenestration density, glomerular hydraulic permeability and GFR and increased fenestration width

We determined glomerular ultrastructural changes focusing on GEnC fenestrations and also glomerular functional measurements, including glomerular ultrafiltration coefficient and GFR in BTBR *ob/ob* mice. The BTBR *ob*^*-*^*/ob*^*-*^ mice at 12 to 20 weeks of age demonstrated increased body weight, hyperglycaemia and albuminuria, compared to litter mate control mice, confirming development of diabetes and diabetic nephropathy (Figure 1A, B and C). Diabetic mice at 20 weeks had significantly decreased fenestration density (Figure 1E) and increased width (Figure 1F) compared to control mice, confirming GEnC fenestral ultrastructural changes.

**Figure 1:**
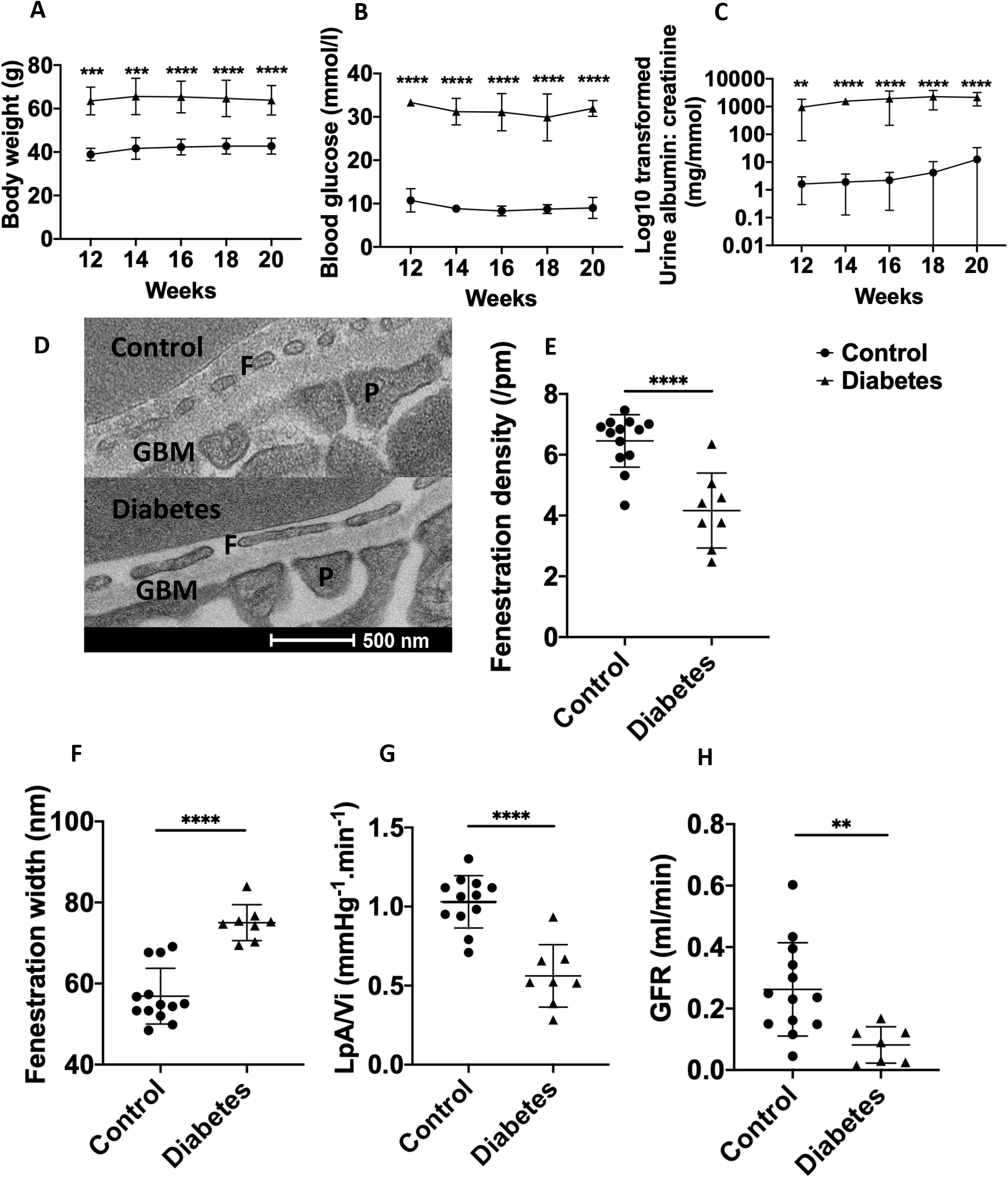
Diabetic nephropathy in BTBR *ob*^*-*^*/ob*^*-*^ mice is associated with decreased fenestration density, glomerular hydraulic permeability and GFR and increased fenestration width. Body weight (A), blood glucose (B) and uACR (C) in BTBR *ob*^*-*^*/ob*^*-*^ mice (n=8) compared to litter mate control mice (BTBR *ob*^*+*^*/ob*^*+*^; n=13). Bodyweight, blood glucose and uACR was significantly increased in BTBR *ob*^*+*^*/ob*^*+*^ at all time points confirming development of diabetes and diabetic nephropathy. Representative transmission electron micrograph demonstrating loss of fenestration density in diabetic mice aged 20 weeks (D). At 20 weeks of age fenestration density (E) was significantly decreased and fenestration width increased (F) in diabetic (n=8) compared to control mice (n=13). Glomerular ultrafiltration coefficient (G; L_P_A/V_i_) was also significantly decreased in diabetic compared to control mice. GFR (H) was significantly decreased in diabetic (n=7) compared to control mice (n=13). ns, not significant; ** P <0.01; *** P <0.001; **** P <0.0001.

Glomerular ultrafiltration coefficient (Figure 1G) and GFR (Figure 1H) were significantly decreased in diabetic compared to control mice, confirming loss of renal filtration function. Podocyte slit density was significantly decreased in diabetic compared to control mice whilst podocyte foot process width and glomerular basement membrane thickness were significantly increased (Supplementary information S1), consistent with development of diabetic nephropathy.

### Fenestration changes are present from 10 weeks of age in BTBR *ob*^*-*^*/ob*^*-*^ mice

We further examined the age at which GEnC fenestration ultrastructural changes developed in BTBR *ob*^*-*^*/ob*^*-*^ mice. GEnC fenestration density was significantly decreased (Figure 2A,B) whilst GEnC fenestration width was significantly increased (Figure 2A,C) in diabetic compared to litter mate control mice aged 10, 15 and 20 weeks but not 6 weeks.

**Figure 2:**
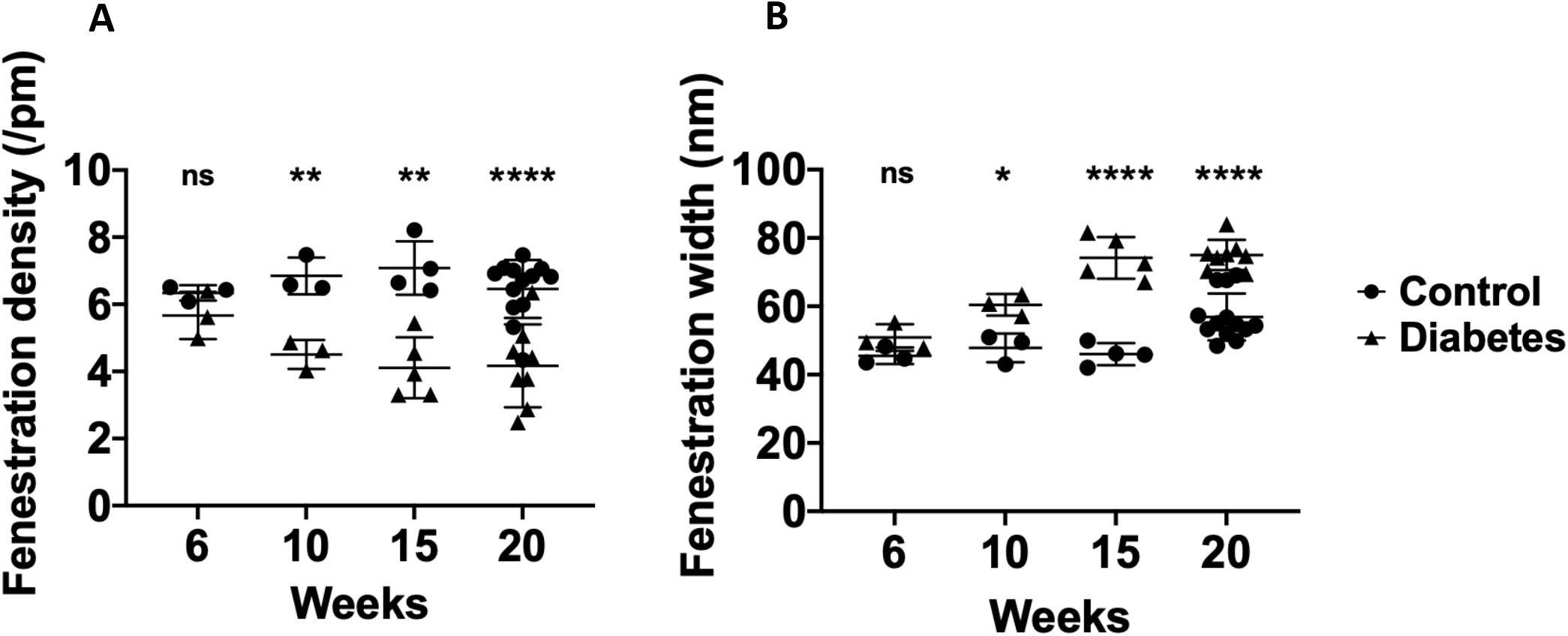
Fenestration changes are present from 10 weeks of age in BTBR *ob*^*-*^*/ob*^*-*^ mice. Representative transmission electron micrograph demonstrating loss of GEnC fenestration density and increasing width in diabetic mice compared to control mice aged 20 weeks (A). GEnC fenestration density was significantly decreased in diabetic compared to litter mate control mice aged 10, 15 and 20 weeks but not 6 weeks (B). GEnC fenestration width was significantly increased in diabetic compared to litter mate control mice aged 10, 15 and 20 weeks but not 6 weeks (C). F, GEnC fenestration; GBM, glomerular basement membrane; P, podocyte foot process; ns, not significant; ** P < 0.01; **** P < 0.0001.

### Reduction in GEnC fenestration density and increase in width are associated with reduced glomerular ultrafiltration coefficient and GFR in BTBR *ob*^*-*^*/ob*^*-*^ mice

We evaluated the glomerular structural and functional relationships to determine the contribution that GEnC fenestrations make to filtration function. Pearson’s correlations (Table 2) revealed significant positive correlations between GEnC fenestration density and both glomerular ultrafiltration coefficient and GFR. Unexpectedly, we identified significant negative correlations between GEnC fenestration width and both glomerular ultrafiltration coefficient and GFR. There were additional significant positive correlations between podocyte slit width and uACR and GBM thickness and uACR (Table 2). There were significant negative correlations between podocyte slit density and uACR and GBM width and glomerular ultrafiltration coefficient (Table 2). Relationships between GEnC fenestral measurements and glomerular functional measurements were further examined by performing univariable linear regression and determining the coefficient of determination (r^2^). There was a significant positive relationship between GEnC fenestration density and both glomerular ultrafiltration coefficient (r^2^ = 0.25, P = 0.019; Supplementary information S2 A) and GFR (r^2^ = 0.27, P = 0.020; Supplementary information S2 B). There was a significant negative relationship between GEnC width and both glomerular ultrafiltration coefficient (r^2^ = 0.33, P = 0.007; Supplementary information S2 C) and GFR (r^2^ = 21, P = 0.041; Supplementary information S2 D).

**Table 2:** Pearson’s correlation between glomerular ultrastructural and functional measurements. GEnC, glomerular endothelial cell; LpA/Vi, Glomerular ultrafiltration coefficient; GFR, glomerular filtration rate; uACR, urinary albumin creatinine ratio; * P < 0.05; ** P < 0.01.

|  | LpA/Vi | GFR | uACR |
| --- | --- | --- | --- |
| GEnC fenestration density | 0.50* | 0.52* | -0.19 |
| GEnC fenestration width | -0.57** | -0.46* | 0.41 |
| Podocyte slit density | 0.39 | 0.38 | -0.52* |
| Podocyte slit width | -0.42 | -0.24 | 0.65** |
| Podocyte foot process width | -0.09 | -0.30 | 0.10 |
| Glomerular basement membrane width | -0.53* | -0.35 | 0.54* |

### Filtration surface area is maintained in diabetes and but this is negatively associated with renal filtration function in diabetes in BTBR *ob*^*-*^*/ob*^*-*^ mice

We next examined if loss of GEnC fenestration density resulted in decreased filtration surface area that could contribute to reduced renal filtration function. We found that filtration surface area was maintained in diabetic compared to control mice at 6, 10, 15 and 20 weeks of age (Figure 3A). We postulate that this is due to increased GEnC fenestration width compensating for the loss of GEnC fenestration density in diabetes. There was no significant overall relationship between filtration surface area and glomerular ultrafiltration coefficient (r^2^ = 0.11, P = 0.143) and GFR (r^2^ = 0.00, P = 0.794). However, when the relationship was examined more closely, there was a positive relationship between filtration surface area and both glomerular ultrafiltration coefficient (Figure 3B) and GFR (Figure 3C) in control mice whereas in diabetic mice the relationships were negative (Figure 3A,B).These findings suggested resistance to flow through the GEnC fenestrations in diabetes.

**Figure 3:**
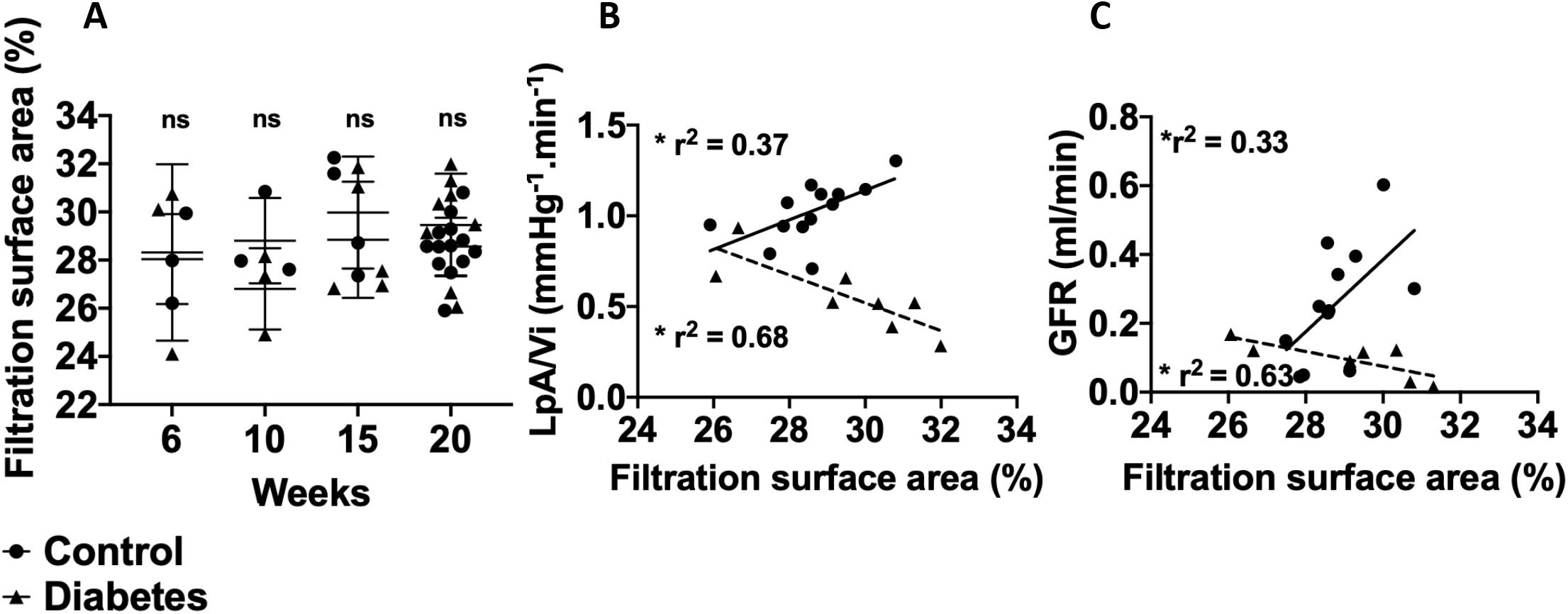
Filtration surface area is maintained in diabetes, but this is negatively associated with renal filtration function in diabetes in BTBR *ob*^*-*^*/ob*^*-*^ mice. Filtration surface area was not significantly different in diabetic compared to control mice aged 6, 10, 15 and 20 weeks (A). There was a significant positive relationship between filtration surface area and glomerular ultrafiltration coefficient (B) and GFR (C) in control mice whereas in diabetic mice the relationships were negative (B,C). Bold line indicates line of regression for control mice (B,C) and dashed line indicates line of regression for diabetic mice (B,C). * P <0.05.

### GEnC fenestrations form diaphragms in diabetic nephropathy and this is negatively associated with renal filtration function

The unexpected finding of a negative association between GEnC fenestration width and both glomerular ultrafiltration coefficient and GFR, and negative relationship between filtration surface area and both glomerular ultrafiltration coefficient and GFR in diabetic mice led us to postulate that this apparent contradiction could be explained by resistance to flow through the GEnC fenestrations by the development of diaphragms in diabetes. We further studied the presence of diaphragms within the GEnC fenestrations as a potential structural impediment to flow. The percentage of diaphragmed fenestrations in diabetic mice significantly increased in diabetic mice being approximately twice that of litter mate control mice (Figure 4 A,B). There was a significant negative relationship between percentage of diaphragmed fenestrations and both glomerular ultrafiltration coefficient (Figure 4C) and GFR (Figure 4D). Furthermore, diaphragmed fenestrations were significantly wider in both diabetic and control mice (Figure 4E). We also demonstrated increased PV1, the only known component of fenestral diaphragms, protein (Figure 4F,G) and mRNA (Figure 4H) expression in glomeruli of diabetic compared to control mice supporting our electron microscopy image findings. Pearson’s correlations revealed significant negative correlations between diaphragmed fenestration width and both glomerular ultrafiltration coefficient (r = 0.56, P = 0.011) and GFR (r = 0.50, P = 0.035). There was no significant correlation between open fenestration width and glomerular ultrafiltration coefficient (r = 0.36, P = 0.113) or GFR (r = 0.10, P = 0.689).

**Figure 4:**
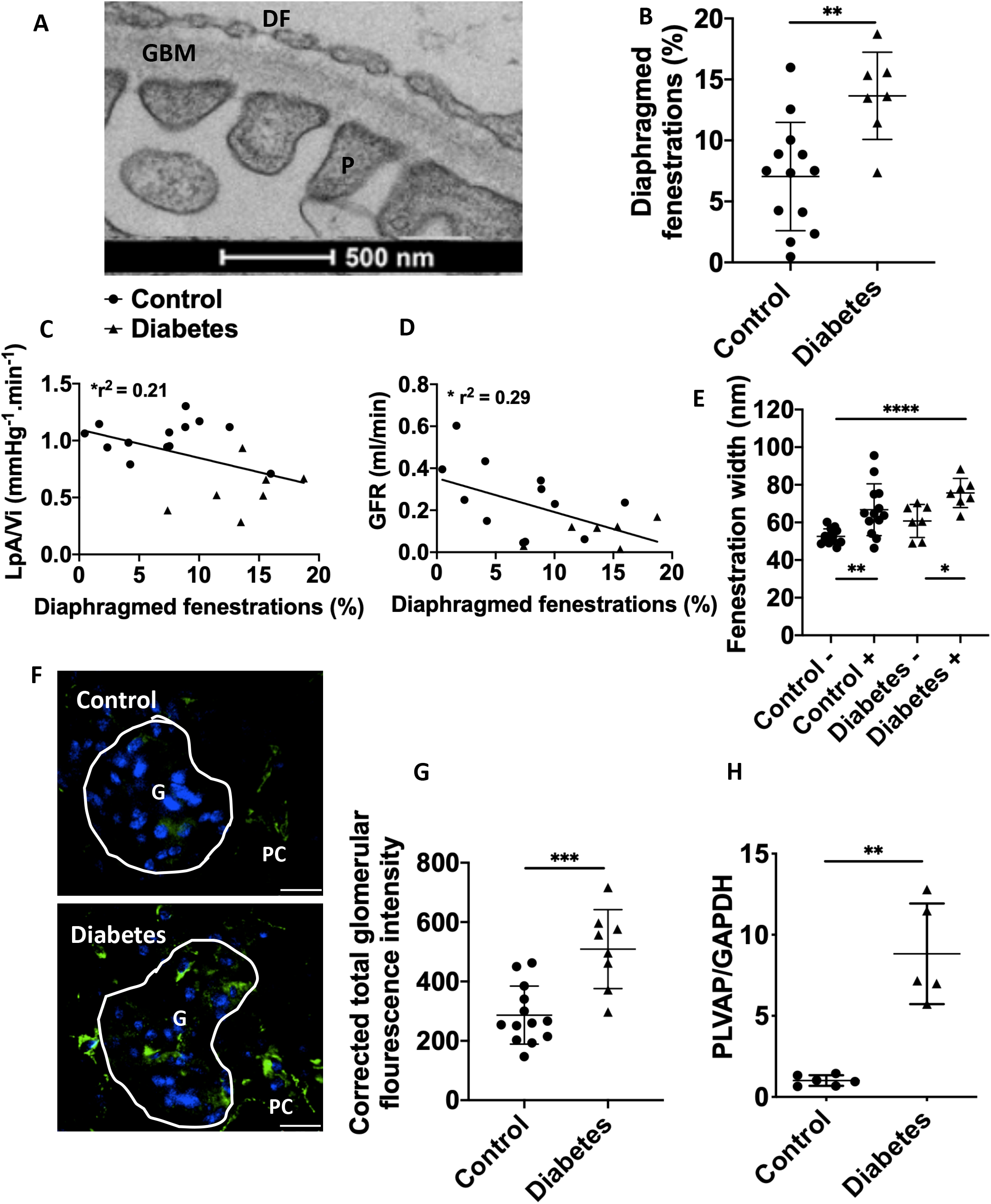
GEnC fenestrations form diaphragms in diabetic nephropathy and this is negatively associated with renal filtration function. Representative TEM image demonstrating the presence of diaphragmed fenestrations in diabetic mice (A). The percentage of diaphragmed fenestrations was significantly increased in diabetic compared to litter mate control mice (B). There was a significant negative relationship between the percentage of diaphragmed fenestrations and both glomerular ultrafiltration coefficient (C) and GFR (D). Fenestration width was significantly higher in diaphragmed fenestrations in both diabetic and control mice (E). Representative image of glomerular PV1 expression determined by immunofluorescence (F). PV1 protein (G) and mRNA (H) glomerular expression was significantly increased in diabetic compared to control mice. Bold line indicates line of regression for all data points in (C, D); Scale bars 25µm (F); DF, diaphragmed fenestration; GBM, glomerular basement membrane; P, podocyte foot process (A); Control -, control open fenestrations; Control +, control diaphragmed fenestrations; Diabetes -, diabetes open fenestrations; Diabetes +, diabetes diaphragmed fenestrations (E); G, glomerulus; PC, peritubular capillary (F); * P <0.05, ** P <0.01, *** P <0.001, *** P <0.0001.

### Eps homology domain proteins 3 (Ehd 3) glomerular expression is decreased in diabetes and is associated with GEnC fenestrations and glomerular ultrafiltration coefficient

Previous studies have demonstrated that Ehd3 and 4 knockout mice demonstrated complete loss of GEnC fenestrations^16^ and that Ehd3 is localised to GEnC fenestrations.^14^ We therefore examined whether Ehd3 expression was altered in diabetes and whether this was associated with GEnC fenestral changes and glomerular ultrafiltration coefficient. Glomerular capillary Ehd3 protein expression, determined by immunohistochemistry, was significantly decreased in diabetic compared to control mice (Figure 5A,B) whilst glomerular Ehd3 mRNA expression was significantly increased (Figure 5C). Ehd3 glomerular capillary protein expression was positively associated with fenestration density (r^2^ = 0.52, P <0.0001) and glomerular ultrafiltration coefficient (r^2^ = 0.59, P <0.001). Ehd4 glomerular protein (Figure 5D) and mRNA (Figure 5E) expression was significantly increased in diabetic compared to control mice, perhaps as a compensatory change to loss of Ehd3 expression. Further, Pearson’s correlation analysis (Table 3) revealed additional positive correlations between Ehd3 and GFR, Ehd4 and GEnC fenestration width and Ehd4 and percentage of diaphragmed fenestrations and negative correlations between Ehd3 and GEnC fenestration width, Ehd3 and percentage of diaphragmed fenestrations, Ehd4 and GEnC fenestration density and Ehd4 and glomerular ultrafiltration coefficient. Podocyte measurements had no significant correlations with Ehd3 and Ehd4 expression except for a positive correlation between podocyte slit density and Ehd3 (Supplementary information S3). GBM width had a significant negative correlation with Ehd3 and Ehd4 (Supplementary information S3).

**Table 3:**
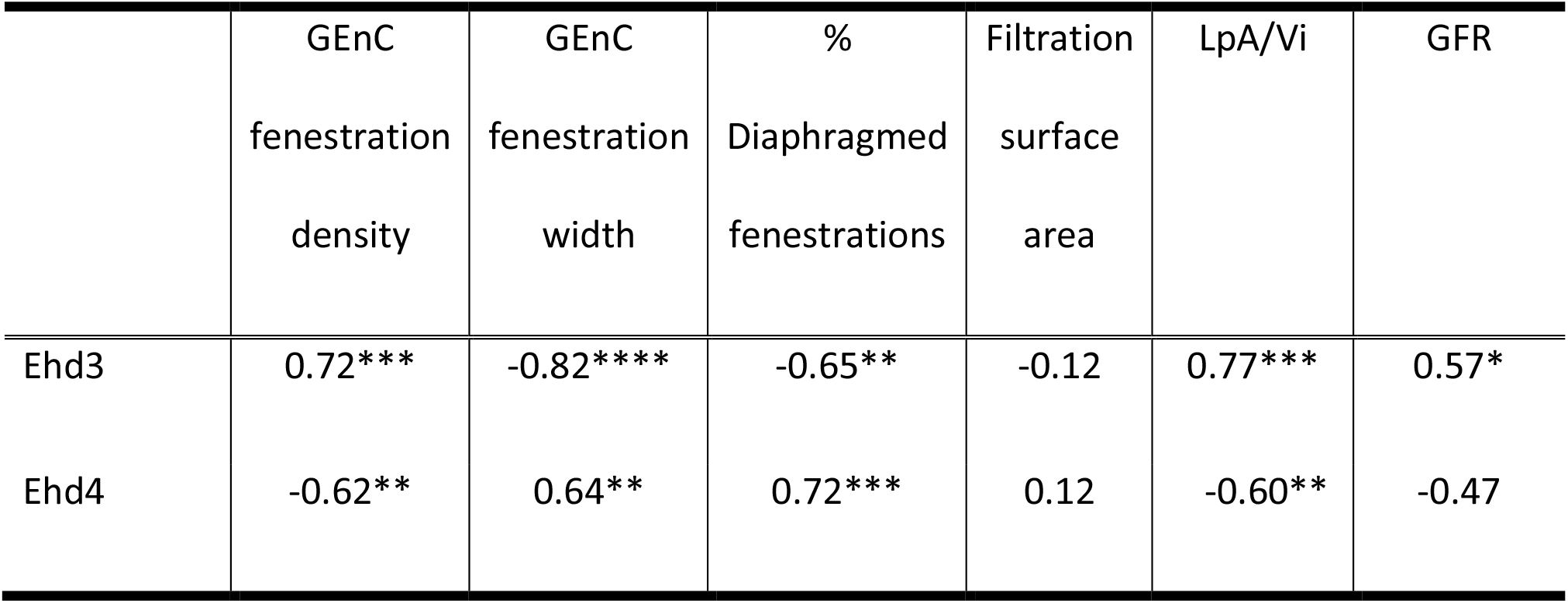
Pearson’s correlation between Ehd3 and Ehd4 expression and GEnC fenestral, filtration surface area and functional measurements. GEnC, glomerular endothelial cell; LpA/Vi, Glomerular ultrafiltration coefficient; GFR, glomerular filtration rate; * P <0.05; ** P <0.01; *** P <0.001; **** P < 0.0001.

|  | GEnC<br>fenestration<br>density | GEnC<br>fenestration<br>width | %<br>Diaphragmed<br>fenestrations | Filtration<br>surface<br>area | LpA/Vi | GFR |
| --- | --- | --- | --- | --- | --- | --- |
| Ehd3 | 0.72*** | -0.82**** | -0.65** | -0.12 | 0.77*** | 0.57* |
| Ehd4 | -0.62** | 0.64** | 0.72*** | 0.12 | -0.60** | -0.47 |

**Figure 5:**
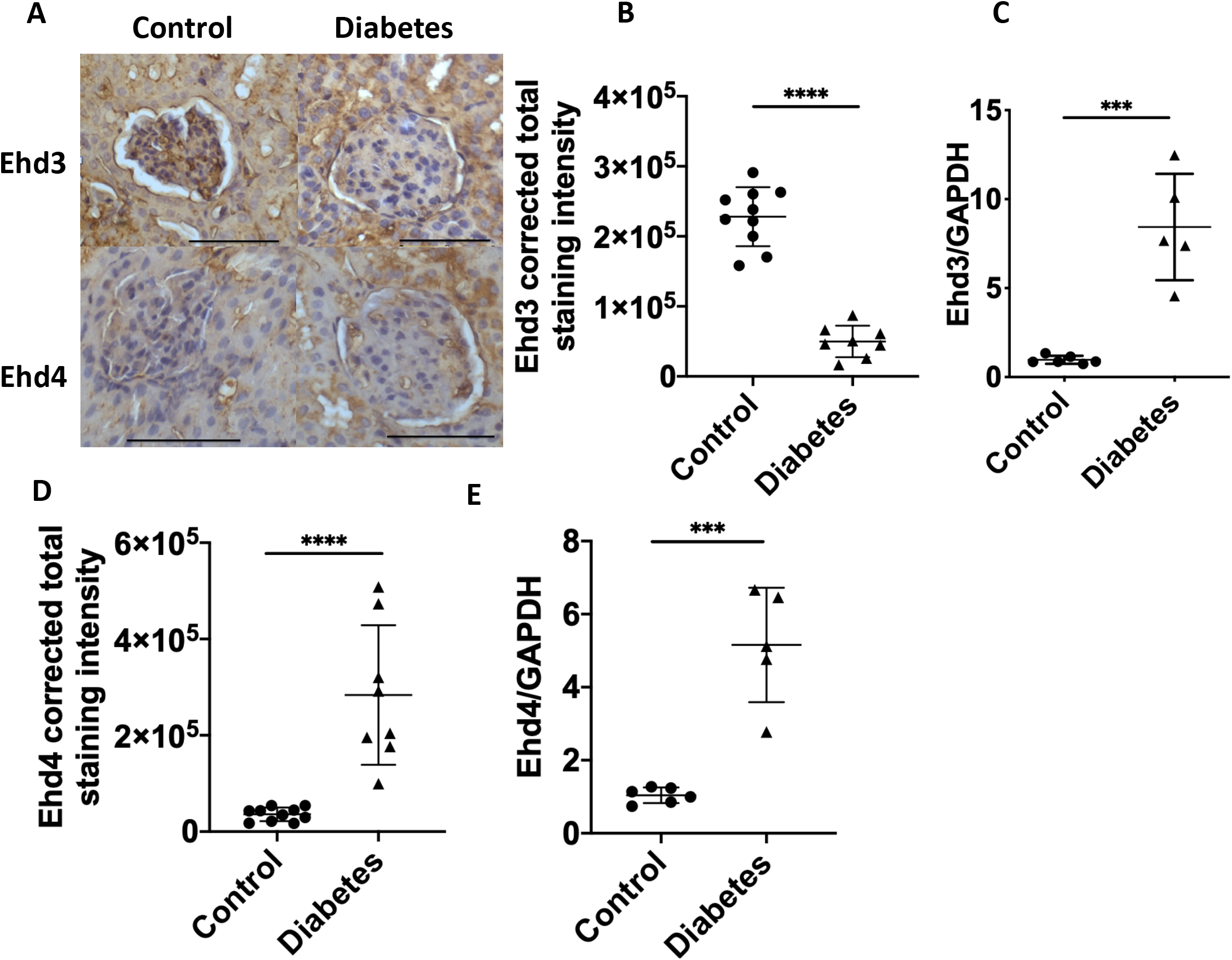
Eps homology domain proteins 3 and 4 (Ehd 3 and 4) expression is altered in diabetes and is associated with GEnC fenestrations and glomerular ultrafiltration coefficient. Representative immunohistochemistry images demonstrating loss of glomerular capillary Ehd3 and increased Ehd4 expression in diabetic mice compared to control mice (A). Glomerular capillary Ehd3 protein expression was significantly decreased (B) and glomerular mRNA expression significantly decreased (C) in diabetic compared to control mice. Glomerular capillary Ehd4 (D) and glomerular mRNA (E) expression was significantly increased in diabetic compared to control mice. Scale bars 100µm (A); * P <0.05, ** P <0.01, *** P <0.001, *** P <0.0001.

#### GEnC fenestral changes are present in human diabetic patients and associated with eGFR and loss of glomerular Ehd3 expression

We finally examined whether our findings identified in diabetic mice were also present in human diabetic patients. GEnC fenestration ultrastructural changes were demonstrated with fenestration density being significantly decreased (Figure 6A) and fenestration width significantly increased (Figure 6B) in human diabetic patients compared to controls. However, in contrast to diabetic mice, the filtration surface area was significantly decreased in diabetic compared to control human patients (Figure 6C). Podocyte slit density was significantly decreased in diabetic compared to control human patients whilst podocyte slit width and glomerular basement membrane thickness were significantly increased (Supplementary information S4) consistent with development of diabetic nephropathy. Pearson’s correlations between glomerular ultrastructural and eGFR measurements (Table 4) revealed significant positive correlations between GEnC fenestration density and eGFR and filtration surface area and eGFR. There were significant negative correlations between GEnC fenestration width and eGFR and GBM thickness and eGFR. There were no significant relationships between podocyte measurements and eGFR. We further studied glomerular Ehd3 expression in diabetic human patients. We identified significantly decreased glomerular capillary Ehd3 expression (Figure 6D,E) in diabetic compared to control human patients. eGFR measurements were available for 4 control and 5 diabetic patients. There was a significant positive relationship between Ehd3 and eGFR (r^2^ = 0.42, P = 0.047). Glomerular capillary Ehd4 expression determined by immunohistochemistry (Supplementary information S5) was significantly increased in in diabetic compared to control human patients considered a potential compensatory change similar to that identified in the diabetic mice.

**Table 4:** Pearson’s correlation between glomerular ultrastructural and eGFR measurements in human patients. eGFR, estimated glomerular filtration rate; * P <0.05; ** P<0.01.

|  | eGFR |
| --- | --- |
| GEnC fenestration density | 0.77** |
| GEnC fenestration width | -0.62* |
| Filtration surface area | 0.71* |
| Podocyte slit density | 0.46 |
| Podocyte slit width | -0.43 |
| Podocyte foot process width | -0.03 |
| Glomerular basement membrane width | -0.83** |

**Figure 6:**
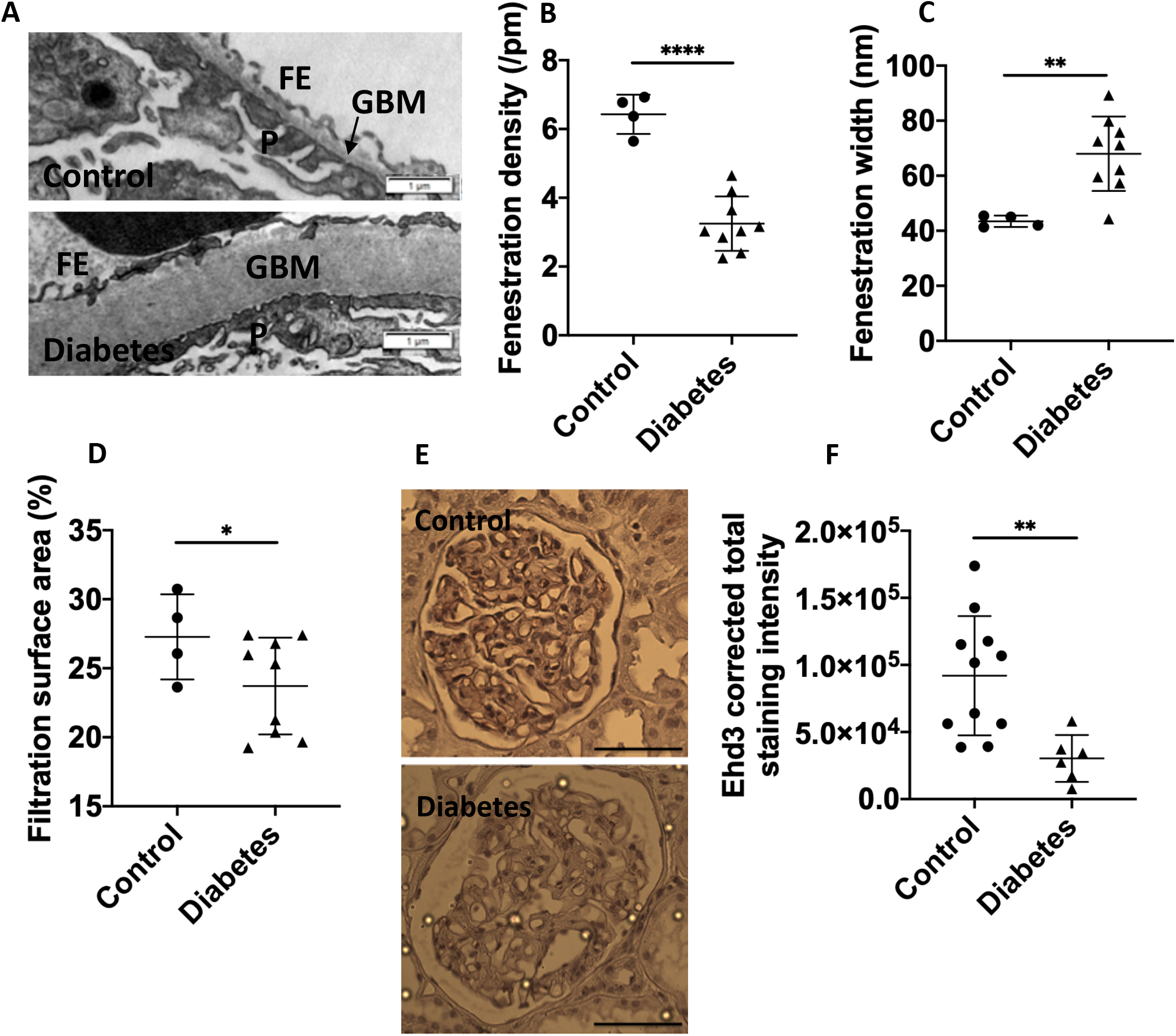
GEnC fenestral changes are present in human diabetic patients and associated with eGFR and loss of glomerular Ehd3 expression. GEnC fenestration density was significantly decreased (A), GEnC fenestration width significantly increased (B) and filtration surface area significantly decreased (C) in human diabetic compared to control patients. Representative immunohistochemistry images demonstrating loss of glomerular capillary Ehd3 (D) in diabetic compared to control human patients. Glomerular capillary Ehd3 protein expression was significantly decreased (E) in diabetic compared to control human patients. Scale bar 100µm (D); * P <0.05; ** P < 0.01; **** P < 0.0001.

Nephroseq datasets were extracted to further examine glomerular PLVAP and Ehd3 expression in additional human diabetic patient cohorts. Glomerular PLVAP expression was studied to evaluate the presence of diaphragmed fenestrations in human patients. PLVAP expression in the Nephroseq ‘Ju CKD Glom’ dataset was examined. There was significantly increased median-centered Log2 PLVAP expression in glomeruli of diabetic compared to control healthy living donor patients (Figure 7A) and a significant negative relationship with eGFR (Figure 7B) suggesting there was increased diaphragmed fenestrations present in human diabetic patients and that this may be contributing to resistance to filtration function. In addition, glomerular Ehd3 expression was studied in the Nephroseq ‘Woroniecka Diabetes Glom’ dataset. There was significantly decreased median-centered Log2 Ehd3 expression in glomeruli of diabetic compared to control healthy living donor patients (Figure 7C) and a significant positive relationship with eGFR (Figure 7D). Further analysis of the Nephroseq database revealed that renal Ehd3 expression was significantly decreased compared to control patients in other kidney diseases (Table 5) including chronic kidney disease and focal segmental glomerulosclerosis.

**Table 5:** Decreased kidney Ehd3 expression (median-centered Log2 expression value) in other kidney diseases compared to control patients. Data extracted from Nephroseq database (www.nephroseq.org). † Nakagawa CKD kidney dataset; †† Hodgin FSGS glom dataset

| Disease comparison | P value | Fold change |
| --- | --- | --- |
| CKD vs normal kidney† | 4.08e<sup>-4</sup> | -11.230 |
| Collapsing Focal Segmental Glomerulosclerosis vs normal kidney†† | 0.003 | -2.285 |

**Figure 7:**
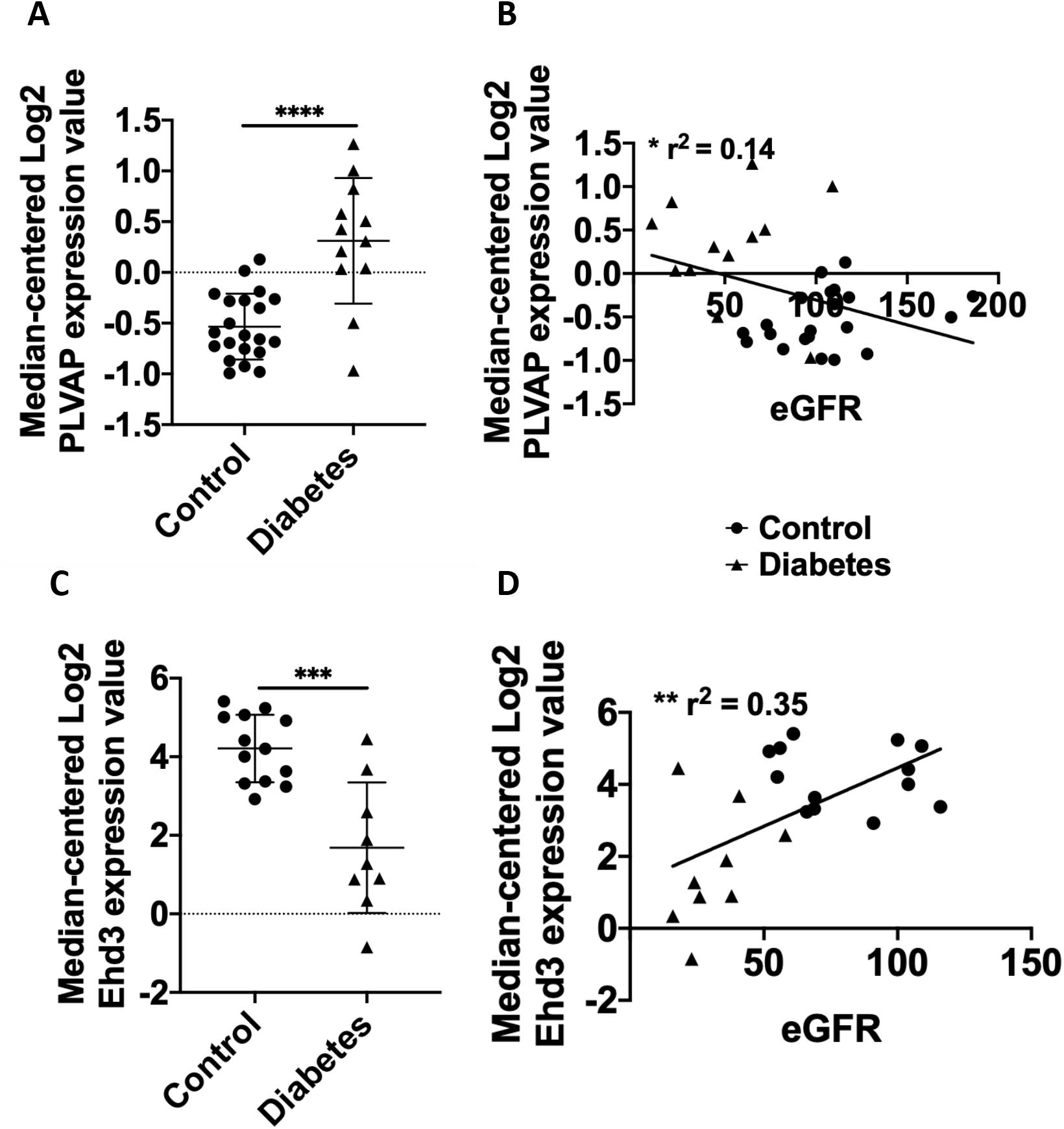
Glomerular PLVAP and Ehd3 expression in Nephroseq datasets. Data from Nephroseq (median-centered Log2 PV1 expression) demonstrating increased PV1 (A) in diabetic (n=12) compared to control healthy living donor patients (n=21) and a significant negative relationship with eGFR (B). Data from Nephroseq (median-centered Log2 Ehd3 expression) demonstrating decreased Ehd3 (C) in diabetic (n=9) compared to control healthy living donor patients (n=13) and a significant positive relationship with eGFR (D). Bold line indicates line of regression for all data points in (C, D); * P <0.05; ** P < 0.01; *** P < 0.001; **** P < 0.0001.

### Ehd3 knockdown decreases fenestration formation in a fenestration forming endothelial cell line

The associations between Ehd3 and GEnC fenestration ultrastructural measurements suggested Ehd3 may directly regulate fenestrations thereby influencing glomerular filtration function. We further examined the role of Ehd3 in regulating fenestrations by knocking down Ehd3 in a fenestration forming cell line (b.END5) (Figure 8 A, B, C). Fenestration formation was significantly decreased in Ehd3 knockdown cells compared to control cells in response to Latrunculin A, an actin depolymerising agent (Figure 8 D, F). Further, there was a complete absence of fenestration formation in Ehd3 knockdown cells compared to control cells in response to mouse VEGF_164_ (Figure 8 D, F). Ehd3 knockdown did not have any significant effect on fenestration width in response to Latrunculin A (Figure 8 E). Fenestrations formed in control cells in response to VEGF_164_ were significantly wider (Figure 8 E, F) and had the appearance of being less organised within sieve plates compared to fenestrations formed in response to Latrunculin A (Figure 8 F).

**Figure 8:**
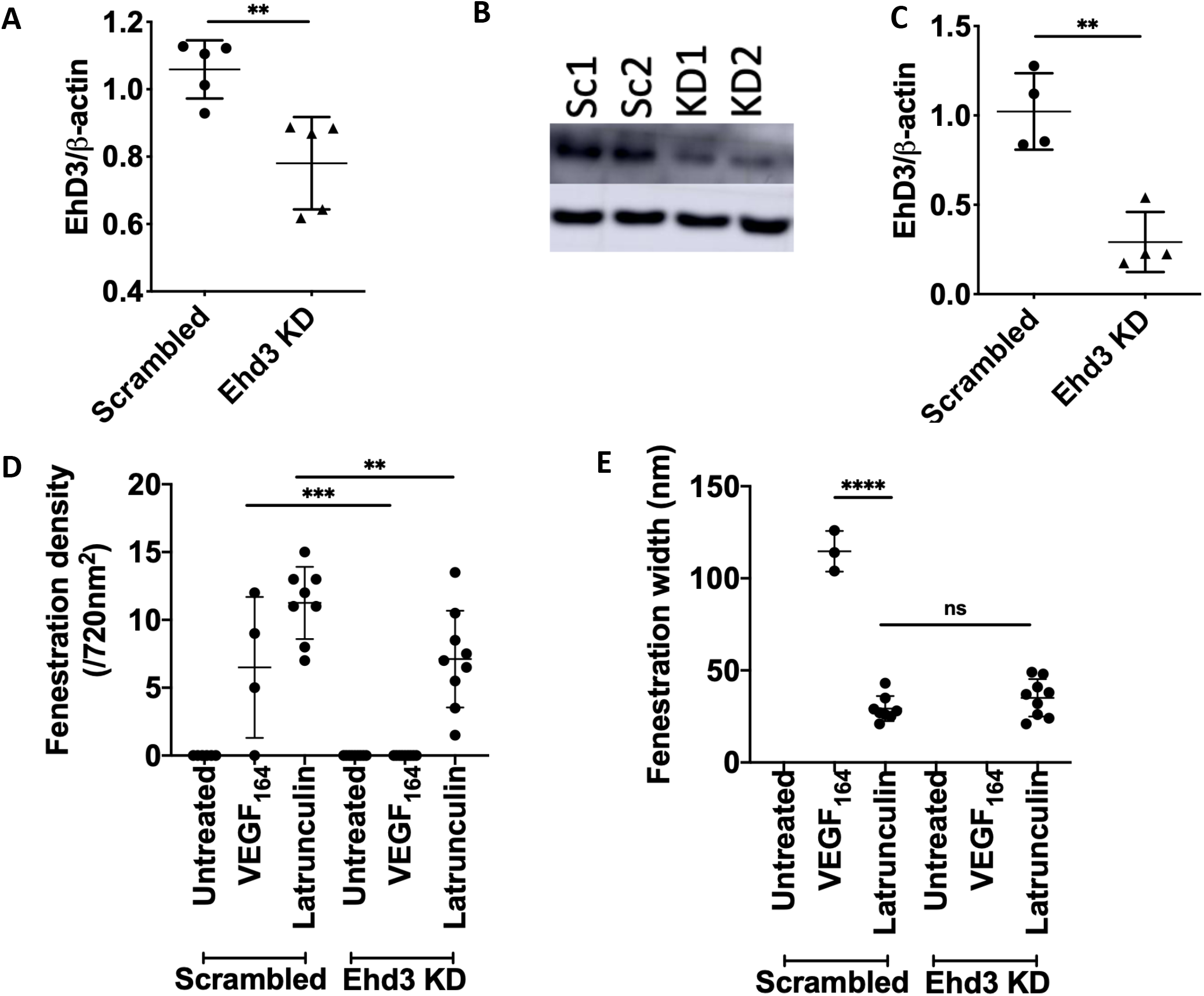
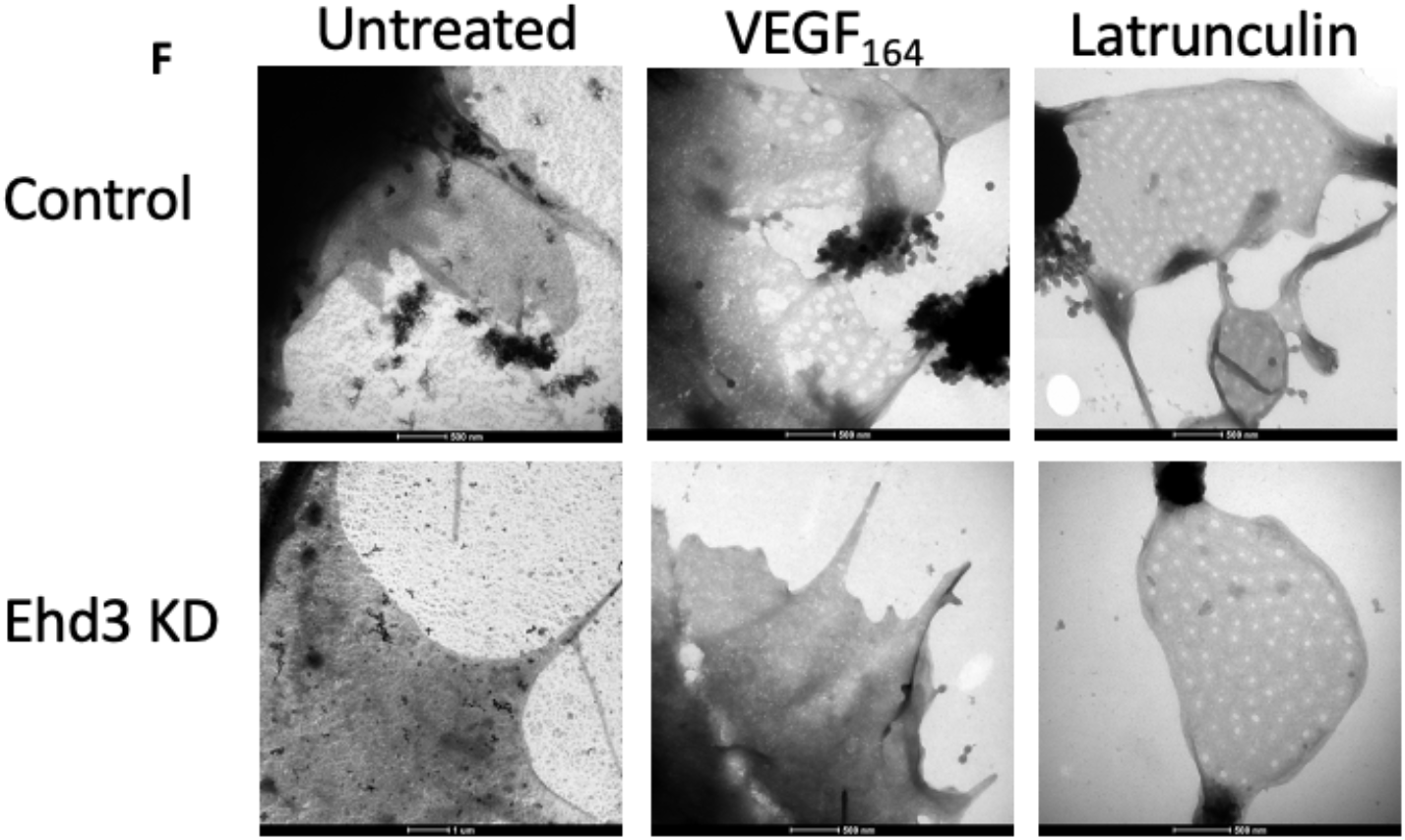
Ehd3 knockdown decreases fenestration formation in a fenestration forming endothelial cell line. Ehd3 mRNA (A) and protein (B, C) knockdown was confirmed in a fenestration forming cell line (mouse brain endothelioma; b.END5). Western blot image confirming knockdown of Ehd3 in b.END5 (C). Fenestration formation was significantly decreased in BEND5 Ehd3 knockdown cells compared to control cells in response to Latrunculin A treatment with complete absence of fenestration formation in response to VEGF_164_ treatment (D). There was no significant difference in fenestration width in response to Latrunculin A treatment in BEND5 Ehd3 cells compared to control cells (E). Fenestrations formed in control cells in response to VEGF_164_ were significantly wider than those formed in response to Latrunculin A treatment (E). Representative whole mount TEM images of BEND5 control and Ehd3 knockdown cells demonstrating fenestration formation in response to VEGF_164_ and Latrunculin A (F). KD, knockdown; Sc, scrambled; ns, not significant; ** P < 0.01; *** P < 0.001; **** P < 0.0001.

## Discussion

Diabetic nephropathy is a significant cause of morbidity and mortality and the healthcare costs associated with it an economic burden. Targeted therapies are urgently needed for patients. In this study, we examine the role of GEnC fenestrations in loss of renal filtration function in diabetic nephropathy. Whilst loss of GEnC fenestrations in diabetes is reported, the full extent of fenestral changes has not been examined. Moreover, the contribution GEnC fenestration changes make to loss of glomerular filtration function in disease and regulatory factors of these structures that may serve as therapeutic targets is unknown. We aimed to comprehensively characterise GEnC ultrastructural changes in diabetic nephropathy in which loss of fenestration density has previously been identified^9^ and examine relationships with renal filtration function. We further aimed to study glomerular Ehd3 expression and *in vitro* loss of Ehd3 as a potential regulator of GEnC fenestrations. We demonstrate that there is decreased GEnC fenestration density, glomerular ultrafiltration coefficient and GFR and increased GEnC fenestration width in a mouse model of diabetic nephropathy and also human diabetic patients. Furthermore, we observed positive relationships between fenestration density and renal filtration measurements, indicating that as fenestrations are lost, renal filtration function declines. The contribution of GEnC fenestration density to renal filtration function likely has relevance to other kidney diseases that are characterised by declining GFR. It is well established that women with pre-eclampsia (a form of thrombotic microangiopathy) have a marked endothelial phenotype with GEnC fenestration loss, and this is considered to contribute to decreased GFR.^17^ In liver sinusoidal endothelial cells, with non-diaphragmed fenestrated endothelium that most closely resembles GEnC fenestrae, fenestration loss is associated with aging^18-20^ and is a proposed mechanism for dyslipidaemia in elderly people. It is therefore possible that age related decline in GFR might also be related to loss of GEnC fenestrations.

Previously, it was suggested, using mathematical modelling, that the contribution of the glomerular endothelium to hydraulic permeability was neglible.^6^ However, the findings of this study contradict this supposition and highlight inaccuracies of mathematical models for predicting physiological measurements. We identified significant relationships between GEnC fenestration measurements and both glomerular ultrafiltration coefficient and GFR. Other structural determinants of hydraulic permeability at the glomerular filtration barrier include the GBM, endothelial junctional integrity, slit diaphragm, slit process width and density and endothelial and epithelial glycocalyx. In the present study, the relationship between podocyte slit width and density and podocyte foot process width and glomerular ultrafiltration coefficient were independently evaluated and were not found to be statistically significant. There was a significant negative relationship between GBM thickness and glomerular ultrafiltration coefficient which is consistent with mathematical modelling predictions. Mathematical modelling predictions have previously suggested that the GBM provides the greatest resistance to glomerular ultrafiltration coefficient contributing to 50% of the resistance.^6^

BTBR *ob*^*-/*^*ob*^*-*^ mice become hyperglycaemic from 3 weeks of age^21^ and albuminuric from 8 weeks of age.^12^ Glomerular structural changes consistent with diabetic nephropathy have been reported to develop in this mouse model by 8 weeks of age.^12^ However, the ultrastructural findings in the GEnC fenestrations have not been described. We demonstrate that GEnC fenestration density is significantly decreased and width significantly increased in BTBR *ob*^*-/*^*ob*^*-*^ compared to control mice by 10 weeks of age. Increased GEnC fenestration width is greater in diabetic rats compared to control rats^22^ consistent with the findings of this study. In the present study, increased GEnC fenestration width with decreasing GEnC fenestration density maintained the GEnC filtration surface area in diabetic mice. In contrast, in human diabetic patients there was a significantly decreased GEnC filtration surface area despite increased GEnC fenestration width. This likely reflects a later stage of disease in the human diabetic patients in which increased GEnC fenestration width cannot compensate for the loss of GEnC fenestration density to maintain filtration surface area. Indeed, mean GEnC fenestration density was lower in human diabetic patients compared to diabetic mice.

Despite maintenance of filtration surface area there was loss of filtration function at the glomerular filtration barrier in diabetic mice. Unexpectedly, we also identified a significant negative relationship between GEnC fenestration width and glomerular ultrafiltration coefficient. We postulated that this may be due to the development of diaphragms in the GEnC fenestrations providing resistance to hydraulic permeability. The number of diaphragmed fenestrae is approximately 4 times greater in GEnC recovering from injury^23^ suggesting a possible role in remodelling. We observed significantly increased diaphragmed fenestrations in diabetic mice in agreement with the findings of a previous study.^24^ Furthermore, we identified increased glomerular PV1, the only known component of diaphragms, expression in diabetic mice and humans. We also observed increased GEnC fenestration width in diaphragmed fenestrations compared to open fenestrations suggesting diaphragms develop in widening GEnC fenestrations. It has been speculated that diaphragmed fenestrations provide some form of structural support or controlled permeability.^25^ The percentage of diaphragmed GEnC fenestrations was found to have a significant negative relationship with glomerular ultrafiltration coefficient and GFR in mice and PV1 expression with eGFR in humans suggesting the presence of diaphragms in GEnC fenestrations does provide resistance to glomerular hydraulic permeability. It has been previously proposed that the fenestral diaphragm does not contribute greatly to resistance to endothelial hydraulic permeability and that the glycocalyx may also account for fenestral resistance.^26^ Endothelial glycocalyx is present within the GEnC fenestrations^27^ although its composition may differ from that on the GEnC luminal surface.^28^ Results of studies describing the glycocalyx on the GEnC luminal surface in diabetes are generally consistent reporting a loss of glyccalyx.^3, 29 30, 31^ It remains unstudied as to whether there is a change in inter-fenestral glycocalyx depth or composition and whether any change may contribute to resistance to glomerular hydraulic permeability.

We demonstrate decreased glomerular capillary Ehd3 protein expression in diabetic mice and human patients. Analysing data from the kidney transcriptomics database Nephroseq, we also identified significantly decreased Ehd3 expression in the glomeruli of diabetic human patients. In the present study, glomerular capillary Ehd3 expression was positively associated with GEnC fenestration density and negatively associated with GEnC fenestration width. Furthermore, loss of Ehd3 expression was associated with loss of glomerular ultrafiltration coefficient and GFR presumably due to aberrations in GEnC fenestrations. We further examined Ehd3 expression using Nephroseq datasets and identified decreased Ehd3 in other kidney diseases including chronic kidney disease (CKD). It is therefore possible that loss of glomerular Ehd3 expression may contribute to GEnC fenestral changes, resulting in decreased glomerular ultrafiltration coefficient, in other kidney diseases characterised by declining GFR. We demonstrated increased glomerular capillary Ehd4 protein expression in diabetic mice and human patients. An increase in glomerular capillary Ehd4 expression has been demonstrated in Ehd3 knockout mice.^16^ Increased Ehd4 expression has also been demonstrated in skeletal muscle in Ehd1 knockout mice.^32^ This suggests increases in Ehd paralogs may be a possible compensatory mechanism for decreased expression of another. Both knockdown of Ehd3 and 4 is required to see a marked glomerular endothelial phenotype with absence of fenestrations.^16^ A quantitative comparison of fenestration density or width in Ehd3^-/-^/Ehd4^-/-^ and Ehd3^-/-^/Ehd4^+/+^ mice was not undertaken in this previous study.^16^

Due to the role of Ehd3 and 4 in endocytic trafficking, it has been postulated that these proteins may regulate recycling and membrane availability of vascular endothelial growth factor receptor 2 (VEGFR2) resulting in altered VEGF signaling.^16^ Indeed, altered VEGFR2 expression was demonstrated in the glomerular endothelium of Ehd3^-/-^/Ehd4^-/-^ mice.^16^ VEGF-A has a critical role in GEnC fenestration regulation. Mice with podocyte specific VEGF-A knockout demonstrate decreased fenestrations^33, 34^ whilst overexpression of the antiangiogenic isoform, VEGF_165_b, also decreases fenestration density and width.^5^ Therefore, it seems plausible that VEGFR2 recycling to the GEnC surface may be altered with decreased Ehd3 expression resulting in loss of VEGF-A mediated maintenance of fenestrations. Ehd4 expression may be increased as a compensatory mechanism to maintain normal recycling. This may be achieved to some degree at early stages of disease however, as Ehd3 expression further declines, endocytic recycling of VEGFR2 becomes deranged. It is also possible that Ehd3 and 4 regulate GEnC fenestrations via other mechanisms for example indirectly via endocytic recycling of other angiogenic factors or directly via interactions with the cellular cytoskeleton. However, Ehd3 does not associate with actin microfilaments and treatment with cytochalasin D, an actin polymerising inhibitor, does not result in altered Ehd3 cellular location.^35^ This suggests it is more likely that Ehd3 indirectly regulates fenestrations via mechanisms related to endocytic recycling.

In summary, we have demonstrated loss of GEnC fenestration density which is associated with decreased glomerular ultrafiltration coefficient and GFR in diabetic nephropathy. GEnC fenestration width increased, an ultrastructural change that may occur to maintain filtration surface area. However, GEnC fenestration width was negatively associated with glomerular ultrafiltration coefficient and GFR considered to be a result of the development of diaphragms in widening fenestrations. There is decreased glomerular Ehd3 expression in diabetes and its association with GEnC fenestration measurements suggests it may play a role in regulating fenestrations. Loss of glomerular Ehd3 expression in disease may contribute to declining glomerular filtration function through aberrant GEnC fenestration regulation. These results suggest that targeted therapies to restore GEnC fenestrations potentially by manipulating glomerular Ehd3 expression may offer potential for restoring renal filtration function in diabetic nephropathy as well as other chronic kidney diseases characterised by similar aberrations in GEnC fenestration regulation and loss of glomerular Ehd3 expression.

## Data Availability

Please contact the first author regarding data availability

## Author contributions

N.C.F, G.W, R.F and S.S designed the study, N.C.F and S.F carried out the experiments, M.B, V.L, D.S and S.H contributed to tissue samples, H.B provided Ehd3 and 4 antibodies, N.C.F, C.N, A.L, M.S and L.D contributed to data analysis, N.C.F, S.F, C.N, M.B, H.M, D.S, GW, R.F and S.S contributed to data interpretation, N.C.F, S.F and C.N created the figures, N.C.F, G.W, R.F and S.S drafted and revised the manuscript; All authors approved the final version of the manuscript.

Acknowledgements

The authors would like to acknowledge the Wolfson Bioimaging Facility, University of Bristol for providing imaging support, Translational Vision Research Laboratory, University College London for training in inducing fenestrations in BEND5. N.C.F and this research were funded by a Wellcome Trust Clinical postdoctoral fellowship award (104507/Z/14/Z).

## Disclosures

### Supplementary material table of contents

**Supplementary information S1:** Podocyte and glomerular basement membrane measurements confirming development of diabetic nephropathy in BTBR *ob*^*+*^*/ob*^*+*^ mice.

**Supplementary information S2:** Reduction in GEnC fenestration density and increase in width are associated with reduced glomerular ultrafiltration coefficient and GFR in BTBR *ob*^-^/*ob*^*-*^ mice.

**Supplementary information S3:** Pearson’s correlation between glomerular Ehd3 and Ehd4 expression and podocyte and GBM measurements.

**Supplementary information S4:** Podocyte and glomerular basement membrane measurements confirming developing of diabetic nephropathy in human diabetic patients.

**Supplementary information S5:** Glomerular Ehd4 expression is significantly increased in human diabetic patients compared to controls.

## Notes

### Competing Interest Statement

The authors have declared no competing interest.

### Author Declarations

University of Bristol and Southmead Hospital research ethics committee H0102/45

## References

1. Satchell, SC, Braet, F: Glomerular endothelial cell fenestrations: an integral component of the glomerular filtration barrier. Am J Physiol Renal Physiol, 296: F947–956, 2009.

2. Salmon, AH, Neal, CR, Bates, DO, Harper, SJ: Vascular endothelial growth factor increases the ultrafiltration coefficient in isolated intact Wistar rat glomeruli. J Physiol, 570: 141–156, 2006.

3. Oltean, S, Qiu, Y, Ferguson, JK, Stevens, M, Neal, C, Russell, A, Kaura, A, Arkill, KP, Harris, K, Symonds, C, Lacey, K, Wijeyaratne, L, Gammons, M, Wylie, E, Hulse, RP, Alsop, C, Cope, G, Damodaran, G, Betteridge, KB, Ramnath, R, Satchell, SC, Foster, RR, Ballmer-Hofer, K, Donaldson, LF, Barratt, J, Baelde, HJ, Harper, SJ, Bates, DO, Salmon, AH: Vascular Endothelial Growth Factor-A165b Is Protective and Restores Endothelial Glycocalyx in Diabetic Nephropathy. J Am Soc Nephrol, 26: 1889–1904, 2015.

4. Stevens, M, Neal, CR, Salmon, AHJ, Bates, DO, Harper, SJ, Oltean, S: Vascular Endothelial Growth Factor-A165b Restores Normal Glomerular Water Permeability in a Diphtheria-Toxin Mouse Model of Glomerular Injury. Nephron, 139: 51–62, 2018.

5. Qiu, Y, Ferguson, J, Oltean, S, Neal, CR, Kaura, A, Bevan, H, Wood, E, Sage, LM, Lanati, S, Nowak, DG, Salmon, AH, Bates, D, Harper, SJ: Overexpression of VEGF165b in podocytes reduces glomerular permeability. J Am Soc Nephrol, 21: 1498–1509, 2010.

6. Drumond, MC, Deen, WM: Structural determinants of glomerular hydraulic permeability. Am J Physiol, 266: F1–12, 1994.

7. Lafayette, RA, Druzin, M, Sibley, R, Derby, G, Malik, T, Huie, P, Polhemus, C, Deen, WM, Myers, BD: Nature of glomerular dysfunction in pre-eclampsia. Kidney Int, 54: 1240–1249, 1998.

8. Alicic, RZ, Rooney, MT, Tuttle, KR: Diabetic Kidney Disease: Challenges, Progress, and Possibilities. Clin J Am Soc Nephrol, 12: 2032–2045, 2017.

9. Weil, EJ, Lemley, KV, Mason, CC, Yee, B, Jones, LI, Blouch, K, Lovato, T, Richardson, M, Myers, BD, Nelson, RG: Podocyte detachment and reduced glomerular capillary endothelial fenestration promote kidney disease in type 2 diabetic nephropathy. Kidney Int, 82: 1010–1017, 2012.

10. Fufaa, GD, Weil, EJ, Lemley, KV, Knowler, WC, Brosius, FC, 3rd, Yee, B, Mauer, M, Nelson, RG: Structural Predictors of Loss of Renal Function in American Indians with Type 2 Diabetes. Clin J Am Soc Nephrol, 11: 254–261, 2016.

11. Toyoda, M, Najafian, B, Kim, Y, Caramori, ML, Mauer, M: Podocyte detachment and reduced glomerular capillary endothelial fenestration in human type 1 diabetic nephropathy. Diabetes, 56: 2155–2160, 2007.

12. Hudkins, KL, Pichaiwong, W, Wietecha, T, Kowalewska, J, Banas, MC, Spencer, MW, Muhlfeld, A, Koelling, M, Pippin, JW, Shankland, SJ, Askari, B, Rabaglia, ME, Keller, MP, Attie, AD, Alpers, CE: BTBR Ob/Ob mutant mice model progressive diabetic nephropathy. J Am Soc Nephrol, 21: 1533–1542, 2010.

13. Grant, BD, Caplan, S: Mechanisms of EHD/RME-1 protein function in endocytic transport. Traffic, 9: 2043–2052, 2008.

14. Patrakka, J, Xiao, Z, Nukui, M, Takemoto, M, He, L, Oddsson, A, Perisic, L, Kaukinen, A, Szigyarto, CA, Uhlen, M, Jalanko, H, Betsholtz, C, Tryggvason, K: Expression and subcellular distribution of novel glomerulus-associated proteins dendrin, ehd3, sh2d4a, plekhh2, and 2310066E14Rik. J Am Soc Nephrol, 18: 689–697, 2007.

15. Karaiskos, N, Rahmatollahi, M, Boltengagen, A, Liu, H, Hoehne, M, Rinschen, M, Schermer, B, Benzing, T, Rajewsky, N, Kocks, C, Kann, M, Muller, RU: A Single-Cell Transcriptome Atlas of the Mouse Glomerulus. J Am Soc Nephrol, 29: 2060–2068, 2018.

16. George, M, Rainey, MA, Naramura, M, Foster, KW, Holzapfel, MS, Willoughby, LL, Ying, G, Goswami, RM, Gurumurthy, CB, Band, V, Satchell, SC, Band, H: Renal thrombotic microangiopathy in mice with combined deletion of endocytic recycling regulators EHD3 and EHD4. PLoS One, 6: e17838, 2011.

17. Stillman, IE, Karumanchi, SA: The glomerular injury of preeclampsia. J Am Soc Nephrol, 18: 2281–2284, 2007.

18. Hilmer, SN, Cogger, VC, Fraser, R, McLean, AJ, Sullivan, D, Le Couteur, DG: Age-related changes in the hepatic sinusoidal endothelium impede lipoprotein transfer in the rat. Hepatology, 42: 1349–1354, 2005.

19. Le Couteur, DG, Cogger, VC, Markus, AM, Harvey, PJ, Yin, ZL, Ansselin, AD, McLean, AJ: Pseudocapillarization and associated energy limitation in the aged rat liver. Hepatology, 33: 537–543, 2001.

20. Hunt, NJ, Lockwood, GP, Warren, A, Mao, H, McCourt, PAG, Le Couteur, DG, Cogger, VC: Manipulating fenestrations in young and old liver sinusoidal endothelial cells. American journal of physiology Gastrointestinal and liver physiology, 316: G144–G154, 2019.

21. Lee, VK, Hosking, BM, Holeniewska, J, Kubala, EC, Lundh von Leithner, P, Gardner, PJ, Foxton, RH, Shima, DT: BTBR ob/ob mouse model of type 2 diabetes exhibits early loss of retinal function and retinal inflammation followed by late vascular changes. Diabetologia, 61: 2422–2432, 2018.

22. Paeng, J, Park, J, Um, JE, Nam, BY, Kang, HY, Kim, S, Oh, HJ, Park, JT, Han, SH, Ryu, DR, Yoo, TH, Kang, SW: The locally activated renin-angiotensin system is involved in albumin permeability in glomerular endothelial cells under high glucose conditions. Nephrol Dial Transplant, 32: 61–72, 2017.

23. Ichimura, K, Stan, RV, Kurihara, H, Sakai, T: Glomerular endothelial cells form diaphragms during development and pathologic conditions. J Am Soc Nephrol, 19: 1463–1471, 2008.

24. Chaiciwamongkol, K, Toomsan, Y, Arnanteerakul, T, Kondo, H, Hipkaeo, W: Occurrence of fenestral diaphragms and knots in renal glomerular endothelia of diabetic mutant MafA-/-MafK+ mice as revealed in embedment-free transmission electron microscopy. Microsc Res Tech, 78: 207–212, 2015.

25. Stan, RV, Tse, D, Deharvengt, SJ, Smits, NC, Xu, Y, Luciano, MR, McGarry, CL, Buitendijk, M, Nemani, KV, Elgueta, R, Kobayashi, T, Shipman, SL, Moodie, KL, Daghlian, CP, Ernst, PA, Lee, HK, Suriawinata, AA, Schned, AR, Longnecker, DS, Fiering, SN, Noelle, RJ, Gimi, B, Shworak, NW, Carriere, C: The diaphragms of fenestrated endothelia: gatekeepers of vascular permeability and blood composition. Dev Cell, 23: 1203–1218, 2012.

26. Levick, JR, Smaje, LH: An analysis of the permeability of a fenestra. Microvasc Res, 33: 233–256, 1987.

27. Okada, H, Takemura, G, Suzuki, K, Oda, K, Takada, C, Hotta, Y, Miyazaki, N, Tsujimoto, A, Muraki, I, Ando, Y, Zaikokuji, R, Matsumoto, A, Kitagaki, H, Tamaoki, Y, Usui, T, Doi, T, Yoshida, T, Yoshida, S, Ushikoshi, H, Toyoda, I, Ogura, S: Three-dimensional ultrastructure of capillary endothelial glycocalyx under normal and experimental endotoxemic conditions. Crit Care, 21: 261, 2017.

28. Avasthi, PS, Koshy, V: The anionic matrix at the rat glomerular endothelial surface. Anat Rec, 220: 258–266, 1988.

29. Ramnath, RD, Butler, MJ, Newman, G, Desideri, S, Russell, A, Lay, AC, Neal, CR, Qiu, Y, Fawaz, S, Onions, KL, Gamez, M, Crompton, M, Michie, C, Finch, N, Coward, RJ, Welsh, GI, Foster, RR, Satchell, SC: Blocking matrix metalloproteinase-mediated syndecan-4 shedding restores the endothelial glycocalyx and glomerular filtration barrier function in early diabetic kidney disease. Kidney Int, 97: 951–965, 2020.

30. Jeansson, M, Granqvist, AB, Nystrom, JS, Haraldsson, B: Functional and molecular alterations of the glomerular barrier in long-term diabetes in mice. Diabetologia, 49: 2200–2209, 2006.

31. Nieuwdorp, M, Mooij, HL, Kroon, J, Atasever, B, Spaan, JA, Ince, C, Holleman, F, Diamant, M, Heine, RJ, Hoekstra, JB, Kastelein, JJ, Stroes, ES, Vink, H: Endothelial glycocalyx damage coincides with microalbuminuria in type 1 diabetes. Diabetes, 55: 1127–1132, 2006.

32. Mate, SE, Van Der Meulen, JH, Arya, P, Bhattacharyya, S, Band, H, Hoffman, EP: Eps homology domain endosomal transport proteins differentially localize to the neuromuscular junction. Skelet Muscle, 2: 19, 2012.

33. Eremina, V, Sood, M, Haigh, J, Nagy, A, Lajoie, G, Ferrara, N, Gerber, HP, Kikkawa, Y, Miner, JH, Quaggin, SE: Glomerular-specific alterations of VEGF-A expression lead to distinct congenital and acquired renal diseases. J Clin Invest, 111: 707–716, 2003.

34. Stevens, M, Neal, CR, Salmon, AHJ, Bates, DO, Harper, SJ, Oltean, S: VEGF-A165 b protects against proteinuria in a mouse model with progressive depletion of all endogenous VEGF-A splice isoforms from the kidney. J Physiol, 595: 6281–6298, 2017.

35. Galperin, E, Benjamin, S, Rapaport, D, Rotem-Yehudar, R, Tolchinsky, S, Horowitz, M: EHD3: a protein that resides in recycling tubular and vesicular membrane structures and interacts with EHD1. Traffic, 3: 575–589, 2002.

